# Incident COVID-19 infections before Omicron in the U.S

**DOI:** 10.1101/2024.10.18.24315777

**Authors:** Rachel Lobay, Ajitesh Srivastava, Ryan J. Tibshirani, Daniel J. McDonald

**Affiliations:** Department of Statistics, The University of British Columbia; Department of Computer and Electrical Engineering, University of Southern California; Department of Statistics, The University of California, Berkeley

**Keywords:** COVID-19, SARS-CoV-2, Infections, Deconvolution, Time series, Seroprevalence, Antibody

## Abstract

The timing and magnitude of COVID-19 infections are of interest to the public and to public health, but these are challenging to ascertain due to the volume of undetected asymptomatic cases and reporting delays. Accurate estimates of COVID-19 infections based on finalized data can improve understanding of the pandemic and provide more meaningful quantification of disease patterns and burden. Therefore, we retrospectively estimate daily incident infections for each U.S. state prior to Omicron. To this end, reported COVID-19 cases are deconvolved to their date of infection onset using delay distributions estimated from the CDC line list. Then, a novel serology-driven model is used to scale these deconvolved cases to account for the unreported infections. The resulting infections incorporate variant-specific incubation periods, reinfections, and waning antigenic immunity. They clearly demonstrate that the reported cases fail to reflect the full extent of disease burden in all states. Most notably, infections were severely underreported during the Delta wave, with an estimated reporting rate as low as 6.3% in New Jersey, 7.3% in Maryland, and 8.4% in Nevada. Moreover, in 44 states, fewer than 1/3 of infections appear as cases reports. Therefore, while reported cases offer a convenient proxy of disease burden, they fail to capture the full extent of infections, and can severely underestimate the true disease burden. This retrospective analysis also estimates other important quantities for every state, including variant-specific deconvolved cases, time-varying case ascertainment ratios, and infection-hospitalization ratios.

## 1 Introduction

Reported COVID-19 cases are a staple in tracking the pandemic at varying geographic resolutions (Dong et al., 2020; The New York Times, 2020; The Washington Post, 2020). Yet, for every case that is eventually reported to public health, several infections are likely to have occurred, and likely much earlier. To see why, it is important to understand *whose* cases are being reported and what differentiates them from unreported cases as well as *when* these case reports happen. Figure 1 shows an idealized path of a symptomatic infection that is eventually reported to public health. This figure illustrates a number of sources of bias in the reporting pipeline. For instance, diagnostic testing mainly targets symptomatic individuals; thus, infected individuals exhibiting little to no symptoms are omitted (Centers for Disease Control and Prevention, 2022). In addition, testing practices, availability, and uptake vary temporally and spatially (European Centre for Disease Prevention and Control, 2020; Hitchings et al., 2021; Pitzer et al., 2021). Finally, cases provide a belated view of the pandemic’s progression, because they are subject to delays due to the viral incubation period, the speed and severity of symptom onset, laboratory confirmation, test turnaround times, and eventual submission to public health (Pellis et al., 2021; Washington State Department of Health, 2020). For these reasons, reported cases are lagging indicators of the course of the pandemic. Furthermore, they do not represent the actual number of new infections that occur on any given day based on exposure to the pathogen. Since there was no large-scale surveillance effort in the United States that reliably tracked symptom onset, let alone infection onset, ascertaining the onset of all *infections* is challenging.

**Figure 1:**
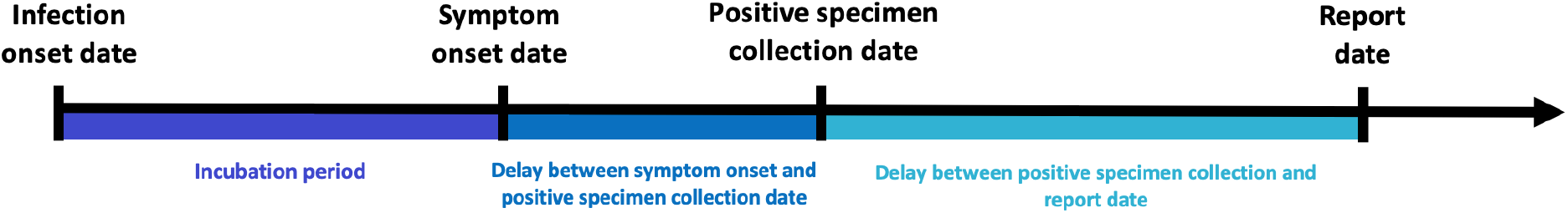
Idealized chain of events from infection onset to case report date for a symptomatic infection that is eventually reported to public health.

Contextualizing the course of the pandemic, understanding the effects of interventions, and drawing insights for future pandemics is challenging because the spatial and temporal behaviour of infections is unknown. While reported cases provide a convenient proxy of the disease burden in a population, it is incomplete, delayed, and misrepresents the true size and timing of the pandemic. Regardless of these difficulties, it is important to the public and to public health to perform a pandemic post-mortem. Estimates of daily incident infections are one such way to measure this and can guide understanding of the pandemic burden over space and time.

In this work, we provide a data-driven reconstruction of daily incident infections for each U.S. state before the onset of Omicron. Using state-level line list data, we estimate state-date specific distributions for the delay from symptom onset to positive specimen date and positive specimen to case report date. We combine these with variant-specific incubation period distributions to deconvolve daily reported COVID-19 cases back to their infection onset, removing the effects of the delays. Finally, we adjust for unreported infections with seroprevalence and reinfection data, accounting for the waning of antigenic immunity over time. A graphical depiction of our procedure is shown in Figure 2. Our results examine features of our infection estimates and the implications of using them, rather than reported cases, to assess the impact of the pandemic. We also produce simple time-varying infection-hospitalization ratios (IHRs) for each state and compare these with case-hospitalization ratios (CHRs). While these analyses provide a glimpse into the utility of our infection estimates, we believe that there is much more to be explored, and we hope that our work serves as a benchmark for future retrospective analyses.

**Figure 2:**
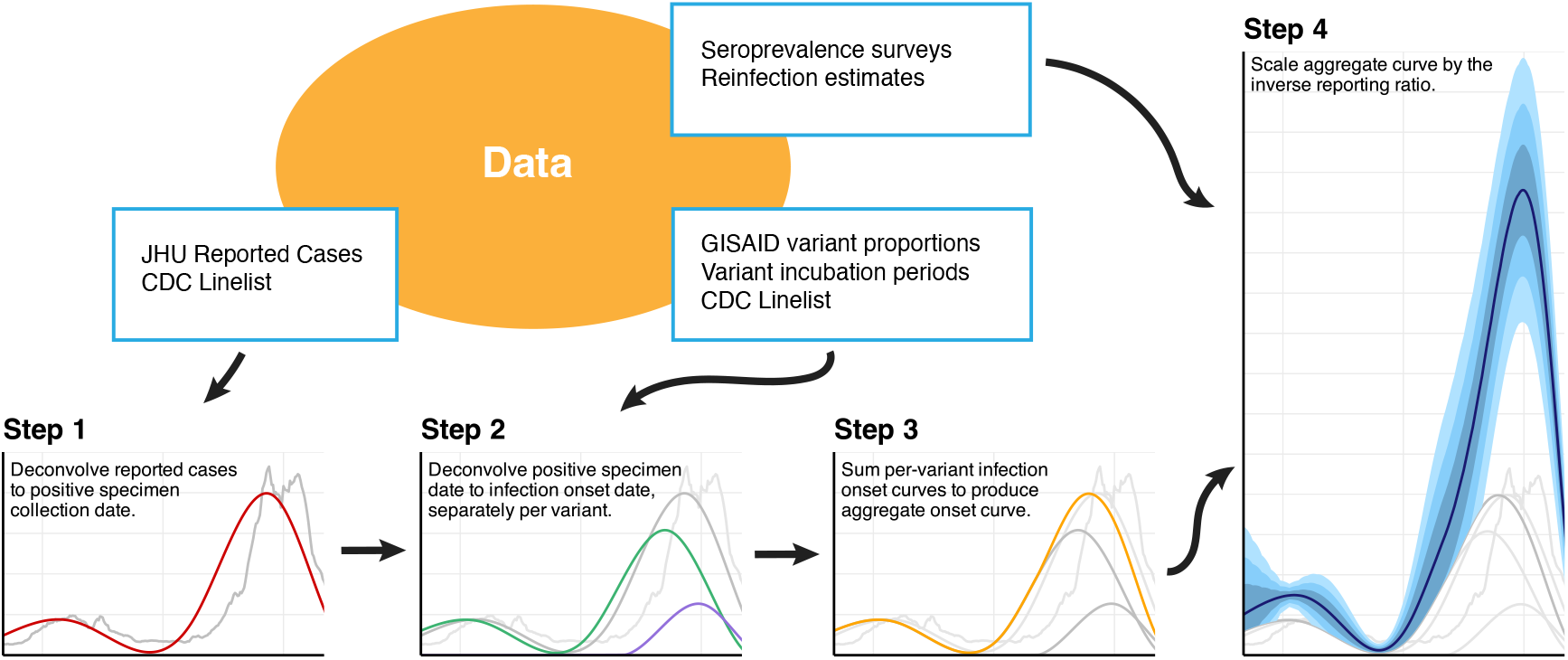
Flowchart of the data and major analysis steps required to get from reported cases to incident infection estimates. In Step 1, we use the CDC line list data to deconvolve reported cases (grey) backward to the date of positive specimen (red). Step 2 separately deconvolves these to the date of infection by variant (Epsilon in Purple, Ancestral in Green), before summing across all variants (orange) in Step 3. Finally, we use seroprevalence survey and time-varying reinfection data to account for the unreported infections.

## 2 Methods

In what follows, we describe how we estimate the daily incident infections for each U.S. state from June 1, 2020 to November 29, 2021. Figure 2 summarizes the major analysis tasks. First, we estimate the delays from positive specimen to report date and use them to push back the reported cases to their sample collection dates. Next, we estimate the delay from symptom onset to sample collection, combine this with variant-specific infection-to-symptom delays, and use these to push back the cases to infection onset. The resulting case estimates are aggregated across variant categories and adjusted by the case ascertainment ratio, estimated with seroprevalence survey data and a model for antigenic immunity.

### 2.1 From reported cases to positive specimen collection

Deconvolution “pushes back” reported cases to the likely date of positive specimen collection. An important aspect of our methods is that deconvolution is not the same as a simple shift, rather it involves the distribution of delays (specific to each state and date). Simply shifting cases back in time would fail to reflect the fact that some cases take much longer to be reported than others (Appendix A).

We will start by describing how the model for deconvolution infers the likely dates of positive specimen collection from reported cases before describing how the CDC line list (Centers for Disease Control and Prevention, 2020a) was used to estimate the necessary delay distributions. Together, these are the ingredients for Step 1 in Figure 2. Define *y*_*ℓ,t*_ to be the number of new cases reported in location *l* at time *t*, as reported by the John Hopkins Center for Systems Science and Engineering (JHU CSSE, Dong et al., 2020) and retrieved with the COVIDcast API (Reinhart et al., 2021). Let *π*_*ℓ,t*_(*k*) be the probability that cases with positive specimen collection at time *t* − *k* are reported at *t*. Then, we model *y*_*ℓ,t*_ as a Gaussian with mean

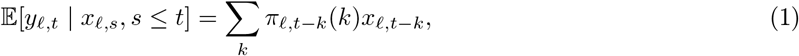

which is a probability weighted sum of the number of positive specimens collected *k* days earlier, *x*_*ℓ,t−k*_. We estimate **x**_*ℓ*_ = {*x*_*ℓ*,1_, …, *x*_*ℓ,T*_} by minimizing the negative log-likelihood with a penalty that encourages smoothness in time. Thus, our estimator is given by

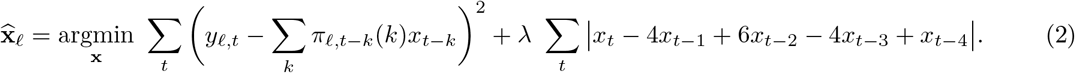

The solution to this minimization problem is an adaptive piecewise cubic polynomial (Tibshirani, 2014, 2022) and can be accurately computed with ease (Jahja et al., 2022; Ramdas and Tibshirani, 2016). We select the tuning parameter *λ* with cross-validation to minimize the out-of-sample reconvolution error.

To estimate the *π*_*ℓ,t*_(*k*) for all states *ℓ*, times *t*, and delays *k*, we use the CDC line list (Centers for Disease Control and Prevention, 2020a). The line list contains three key dates of interest for many cases that eventually appear in case reports: symptom onset, positive specimen collection, and report to the CDC. Handling missingness in these dates requires careful attention (Appendix B). Define *z*_*ℓ,t*_ to be a case report occurring at time *t* in location *ℓ*. We assume that positive samples are reported within 60 days and that no test is reported on the same date as it was collected. Under these assumptions, let *N*_*ℓ,t*_ be the total number of *z*_*ℓ,r*_ with positive specimen collection date *r* in a window *r* ∈ [*t* − 75 + 1, *t* + 60] around *t*. Then, we compute the observed probability mass function (pmf)

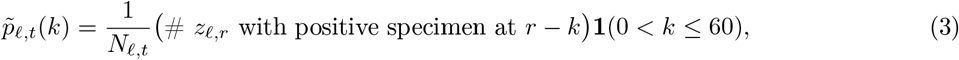

where **1**(*Z*) = 1 if *Z* is true and 0 otherwise. We also compute a similar national pmf, 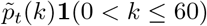, without restricting to location *ℓ*. Next, let *α*_*ℓ,t*_ be the ratio of *N*_*ℓ,t*_ to the number of cases reported by JHU CSSE (Dong et al., 2020) in the window [*t* − 60 + 2, *t* + 75]. Then, compute 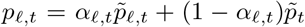. This construction was adopted to allow for more reliance on the state estimate when a larger fraction the JHU cases reports appear in the CDC line list. We calculate the mean *m*_*ℓ,t*_ and variance *v*_*ℓ,t*_ of the pmf {*p*_*ℓ,t*_(*k*)} and estimate the best-fitting gamma distribution by solving the moment equations *m*_*ℓ,t*_ = *α*_*ℓ,t*_*θ*_*ℓ,t*_ and 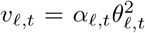 for the shape *α*_*ℓ,t*_ and scale *θ*_*ℓ,t*_. Finally, we discretize the resulting gamma density to the original support to produce an estimate 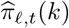 of the delay distribution *π*_*ℓ,t*_(*k*). Additional details are deferred to Appendix C.

### 2.2 From positive specimen collection to infection onset

To continue, pushing positive specimen collection time back to infection onset (Step 2 in Figure 2), we use a procedure very similar to that described above and specified in Equations (1) and (2). However, because the delays involve the time from infection to symptom onset, these must be variant-specific. We use our estimates from Section 2.1, 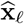, but we weight them corresponding to the mix of variants in circulation. To estimate the daily proportions of the variants circulating in each state, we use GISAID genomic sequencing data from CoVariants.org (Elbe and Buckland-Merrett, 2017; Hodcroft, 2021), and estimate a multinomial logistic regression model. This procedure is now standard (Appendix D) (Annavajhala et al., 2021; Figgins and Bedford, 2021; Obermeyer et al., 2022). The resulting estimated probability of variant *j* is given by 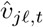.

To estimate variant-specific delays from infection to positive specimen collection, we convolve the location-time-specific symptom-to-test distributions (that are estimated from the CDC line list in the same way as in Section 2.1), with variant-specific incubation periods. The convolution of these yields a distribution 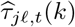. Details on the convolution and its inputs are in Appendices F to H.

Analogous to Equations (1) and (2), for each variant *j*, we model the variant-specific, deconvolved cases as Gaussian with mean

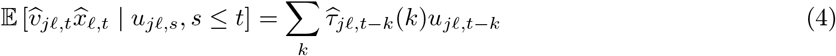

and estimate **u**_*jℓ*_ by minimizing the negative loglikelihood with a penalty to encourage smoothness:

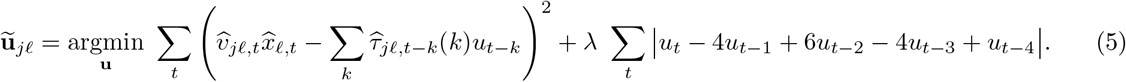

We call the solution 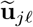 the *variant-specific deconvolved cases* and emphasize that these are cases that will eventually be reported to public health. Because this deconvolution is performed separately for each location and variant, we sum over the variants at each time *t*, and denote the total deconvolved cases at location *ℓ* as 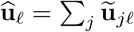 (Step 3 in Figure 2). Note that these deconvolved cases are now indexed by the time of infection onset rather than case report.

### 2.3 Inverse reporting ratio and the antibody prevalence model

To capture the unreported infections, it is necessary to adjust these deconvolved case estimates by the inverse reporting ratio, the ratio of the number of incident infections to incident reported infections (Step 4 in Figure 2). Seroprevalence of anti-nucleocapsid antibodies represents the percentage of people who have at least one resolving or past infection (Centers for Disease Control and Prevention, 2020b), so we develop a model that uses the change in subsequent seroprevalence measurements to estimate all new infections. We use two seroprevalence surveys to estimate the proportion of the population with evidence of previous infection in each state over time (Appendix I) (Centers for Disease Control and Prevention, 2021a,b).

To account for different surveys occurring on different dates with roughly weekly availability and measurement error, we treat actual seroprevalence *s*_*ℓ,m*_ as a latent variable available on Monday (using *m* rather than *t* to denote Mondays). Therefore, the observed seroprevalence survey measurements 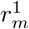 and 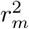 are modelled as Gaussian,

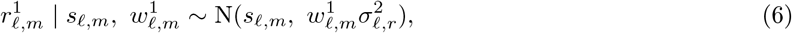

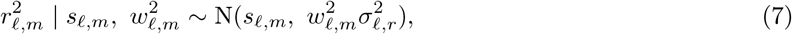

with source-specific measurement errors, 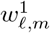 and 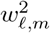, that scale proportional to reported uncertainty.

To complete the model, we suppose that latent seroprevalence is modeled as Gaussian with mean given by a fraction of the previous seroprevalence measurement at *m* plus the reinfection-adjusted deconvolved cases multiplied by the inverse reporting ratio at time *m*:

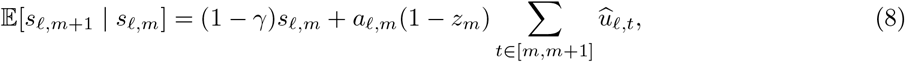

where 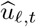 are deconvolved cases (from Section 2.2), *z*_*m*_ is the fraction of reinfections, and *a*_*ℓ,m*_ is the inverse reporting ratio. Note that *γ* is the fraction of people whose level of infection-induced antibodies falls below the detection threshold between time *t* and time *t* + 1. The daily fraction of new infections *z*_*t*_ are based on surveillance work conducted by the Southern Nevada Health District (Ruff et al., 2022), and these estimates are broadly similar to those in other locations with available data (Hawaii Department of Health, 2022; New York State Department of Health, 2023; Ruff et al., 2022; Washington State Department of Health, 2022). Finally, we specify the time-varying evolution of the inverse reporting ratio as Gaussian with expectation,

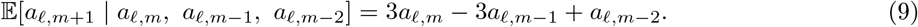

This construction for Equation (9) results in estimates that vary smoothly in time.

The antibody prevalence model specified by Equations (6) to (9) is a state space model with latent variables **s**_*ℓ*_ and **a**_*ℓ*_. In this way, the latent variables and all unknown parameters can be estimated using maximum likelihood, despite missing or irregularly-spaced survey measurements. Additionally, latent quantities can be extrapolated beyond the times of measured seroprevalence. Additional details of this methodology and the computation of the associated uncertainty measurements are in Appendix J.

### 2.4 Lagged correlation to hospitalizations and time-varying IHRs

From the COVIDcast API (Reinhart et al., 2021), we retrieve the daily number of confirmed COVID-19 hospital admissions for each state that are collected by the U.S. Department of Health and Human Services (HHS). We use our infection estimates 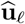 to compute the lagged correlation with hospitalizations. The goal of this analysis is to find the lag between infection and hospitalization rates that gives the highest average rank-based correlation across U.S. states. Thus, we consider a wide range of possible lag values ranging from 1 to 25 days. Then, to assess the impact of our modelling choices, particularly the contribution of the main steps to the lagged correlation analysis, we conduct an ablation study that is detailed in Appendix K.

For each considered lag, we calculate Spearman’s correlation between the state infection and hospitalization rates for each observed between June 1, 2020 to November 29, 2021 with a center-aligned rolling window of 61 days. We then average these correlations across all states and times for each lag.

The lag that leads to the highest average correlation is used to estimate the time-varying IHRs for each state. The IHR is computed by dividing the number of individuals who are hospitalized due to COVID-19 by the estimated total number who were infected on the lagged number of days before. To stabilize these lagged IHR estimates, we average these hospitalizations and infections within a window of 31 days centered on the date of interest, rather than just using one pair of dates for each computation.

## 3 Results

### 3.1 Infection estimates and cases-to-infections ratios across the U.S. states

Prior to Omicron, the largest infection outbreaks were observed in the late summer and early fall of 2021 in Louisiana, Georgia, Idaho, and Montana (Figures 3 to 4). During this time, the state with the highest rate of infections on a single day is Louisiana, with 476 infections per 100K on July 20, 2021. For comparison, the state’s 7-day average case rate peaks at 126 cases per 100K on August 13, 2021. Idaho follows with an infections peak of 457 per 100K on September 7, 2021, and a case peak of 76 per 100K occurring shortly thereafter on September 13, 2021. The period of lowest viral transmission is observed in the summer of 2020, when Vermont has fewer than 10 infections per 100K per week from June to August, the longest such lull observed for any state.

**Figure 3:**
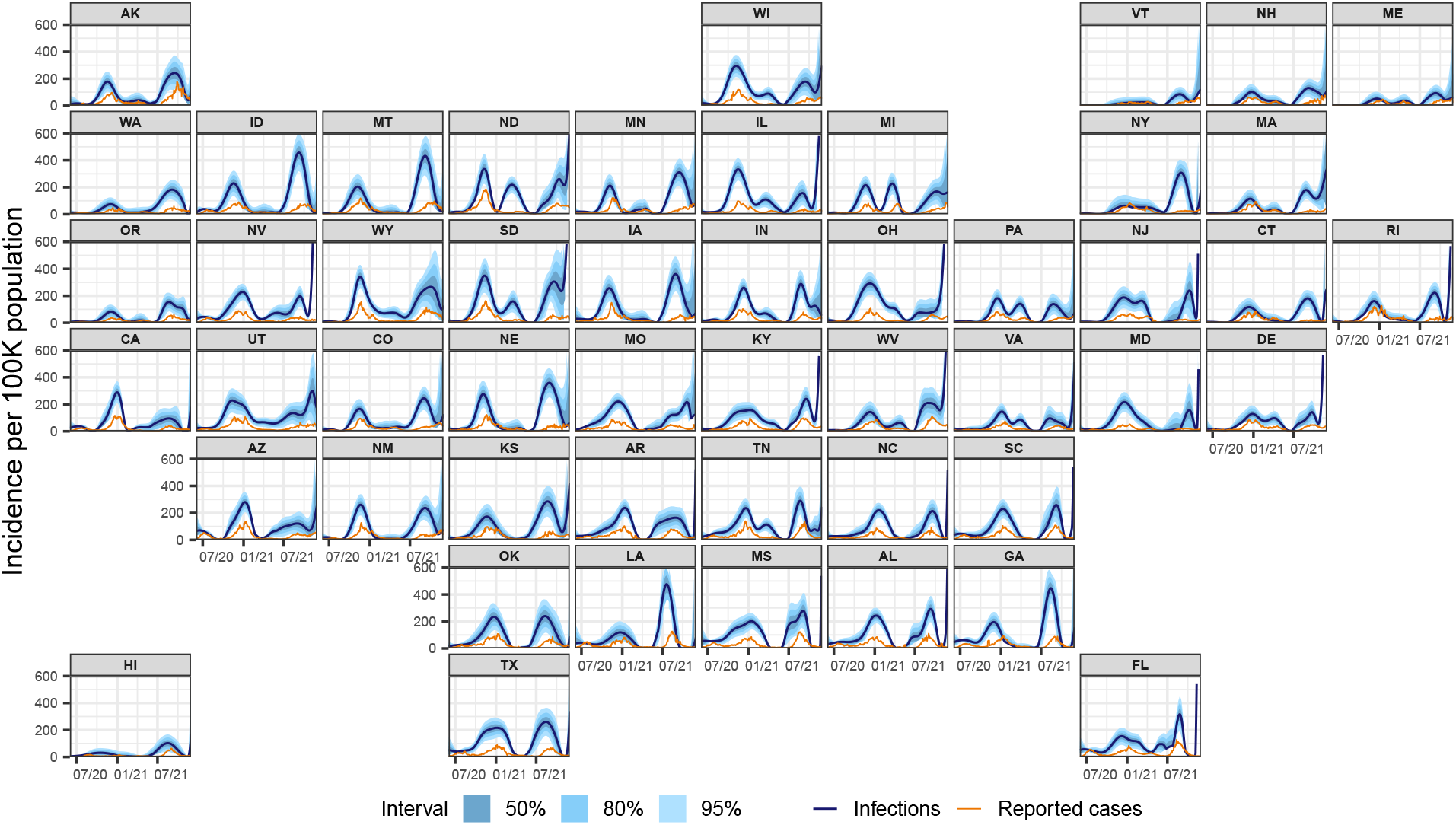
Estimates of the daily new infections per 100,000 population for each U.S. state from June 1, 2020 to November 29, 2021 (dark blue line). The blue shaded regions depict the 50, 80, and 95% intervals for the estimates, while the orange line represents the trailing 7-day average of reported cases per 100,000.

**Figure 4:**
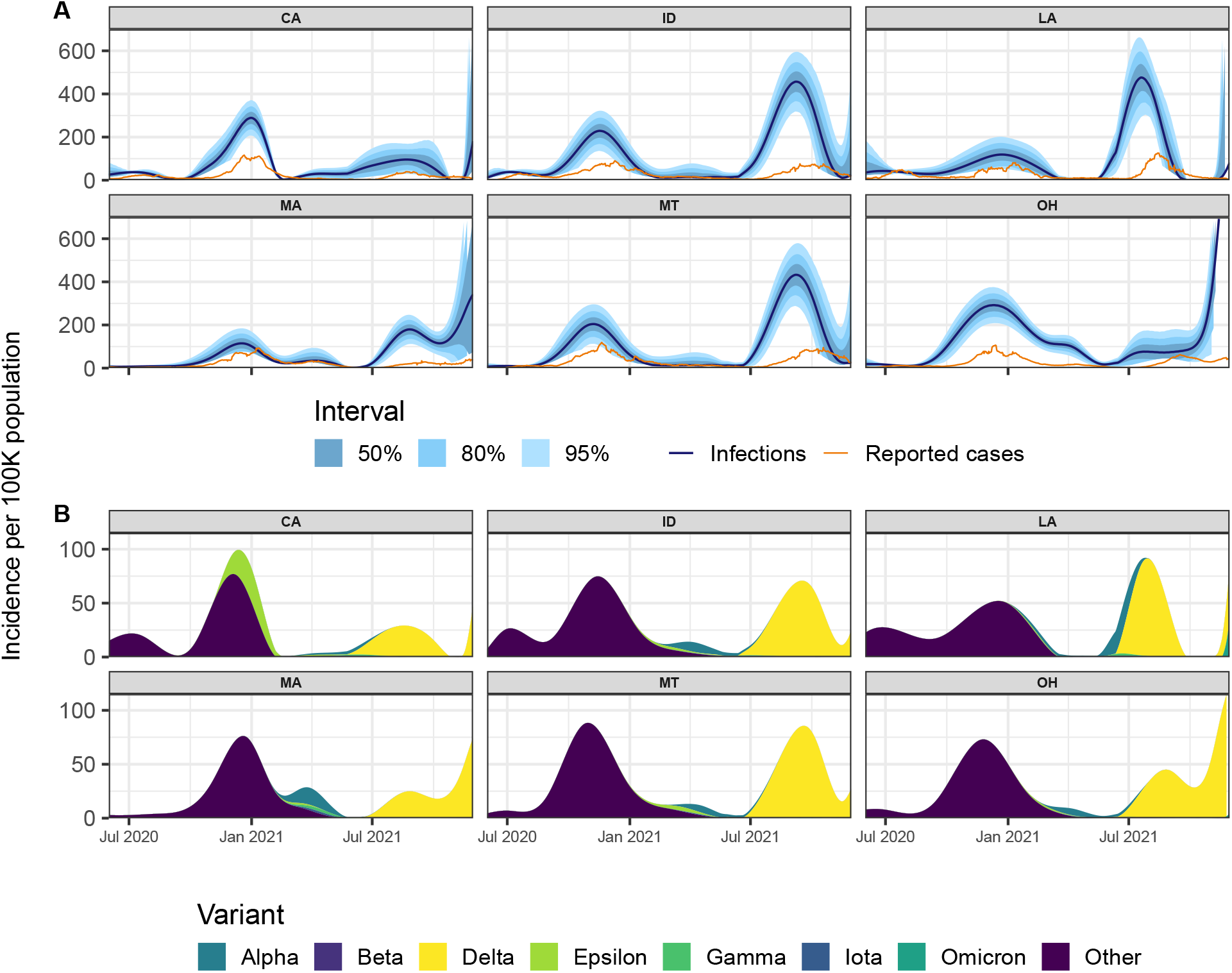
Panel A: Reported cases (orange) and estimates of daily new infections (dark blue) per 100K inhabitants. The blue shaded regions indicate 50, 80, and 95% confidence bands. Panel B: Deconvolved cases colored by variant per 100K inhabitants.

Nearly all states exhibit two major waves in infections—the Ancestral wave began in the fall of 2020 and extended into the winter season, while the Delta wave started in the late summer of 2021 and continued into mid-fall. In general, greater similarities in the strength and magnitude of outbreaks emerge in small clusters of states that border each other (Idaho and Montana; North and South Carolina) present waves of infections that mirror each other in amplitude and timing.

While the Ancestral, Alpha, and Delta waves are visible for most states, there are clear outbreaks in unreported infections that are not easily detectable from cases alone. For example, a wave of infections is evident in North and South Dakota over the spring of 2021 that is virtually undetectable from reported cases. Similarly, in late-summer 2021, the Delta wave is only faintly detectable from cases in a number of Northeastern states, while infections suggest that it has already begun in earnest.

Moreover, cases tend to severely underestimate infections during Delta for many states, more so than in earlier waves (Figure 3). The most extreme was New Jersey, where about 6.3% of estimated infections were eventually reported as cases. Similarly low are Maryland (7.3%), Nevada (8.4%), and South Dakota (10.0%). In 44 states, fewer than 1/3 of infections eventually appear in case reports. The cases-to-infections ratio was larger in earlier waves, and its effects were most apparent in different regions. During Alpha, Louisiana had the lowest ratio of infections to cases (11.9%) followed by California (13.6%). Such patterns are less apparent during the Ancestral wave, where Ohio and Maryland had the lowest ratio of reported cases to infections at 21.4% and 21.7%, respectively.

Figure 5 shows that using cases as a proxy for infections can lead to misunderstandings in the locations that are affected and the extent to which they are affected. For example, on October 20, 2020, while case rates are elevated in a handful of upper-Midwestern states (namely, North and South Dakota), infection rates are elevated to a similar extent in the surrounding states as well, indicating a wider impact than suggested by cases alone. On July 20, 2021, while the map of case rates shows low and geographically consistent impact, infection rates reveal that Texas, Louisiana, Georgia, and their neighbors are hotspots.

**Figure 5:**
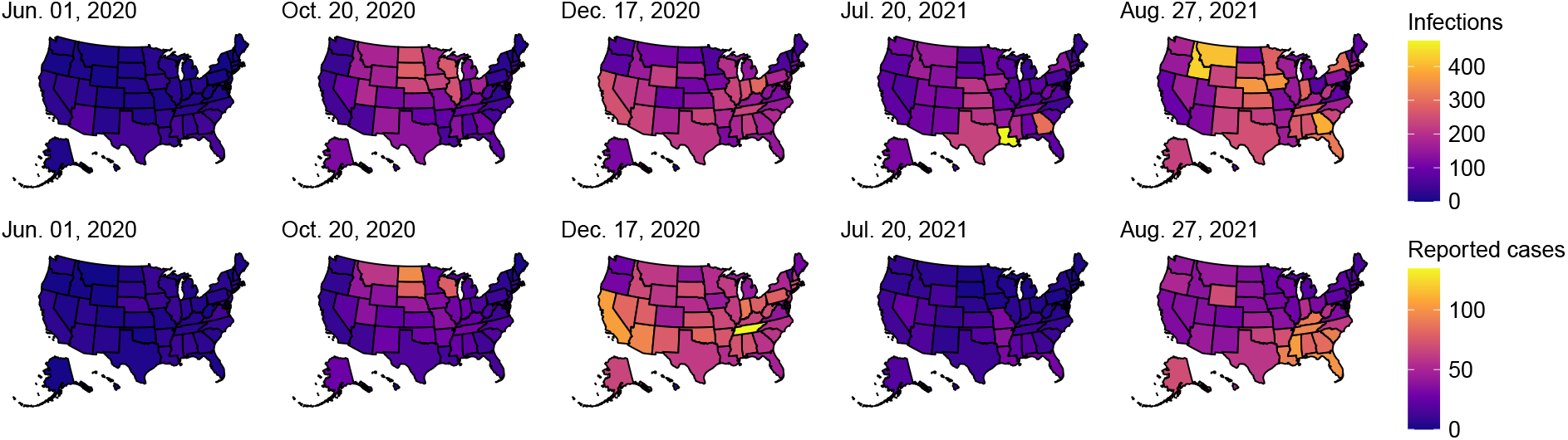
Choropleth maps of the state-level estimates of the daily new infections per 100K (top row) and the daily new cases per 100K (bottom row) for five select dates between June 1, 2020 and November 29, 2021. Note that the first date was chosen as a baseline, while the other dates were chosen because they present large counts of infections across all states. In particular, the third and fifth dates present the largest number of total infections across the 50 states within those calendar years.

By focusing on states with elevated cases, infection outbreaks may be overlooked. For instance, on August 27, 2021, Montana and Idaho have some of the highest infection rates (Figure 5). In contrast, their case rates are unremarkable (the highest case rates tend to be in the Southeast). Infection outbreaks tend to precede case outbreaks, though the lead time can vary widely. During the Delta wave, infections in Montana peaked about 41 days before cases, while in Idaho, they peaked about 6 days before cases (Figure 3). During the Ancestral wave, infections peaked about 12 days earlier than cases in Montana and 24 days earlier in Idaho, demonstrating a notable shift in lead times.

### 3.2 Insights from cross-correlations, IHRs and CHRs

The maximum Spearman’s correlation between infections and hospitalizations is 0.48 and occurs at a lag of 13 days (Figure 6). In contrast, we find that the largest average Spearman correlation for cases is 0.69 and occurs at a lag of 1 day. That is, case reports are nearly contemporaneous to hospitalizations, while infection estimates clearly precede them.

**Figure 6:**
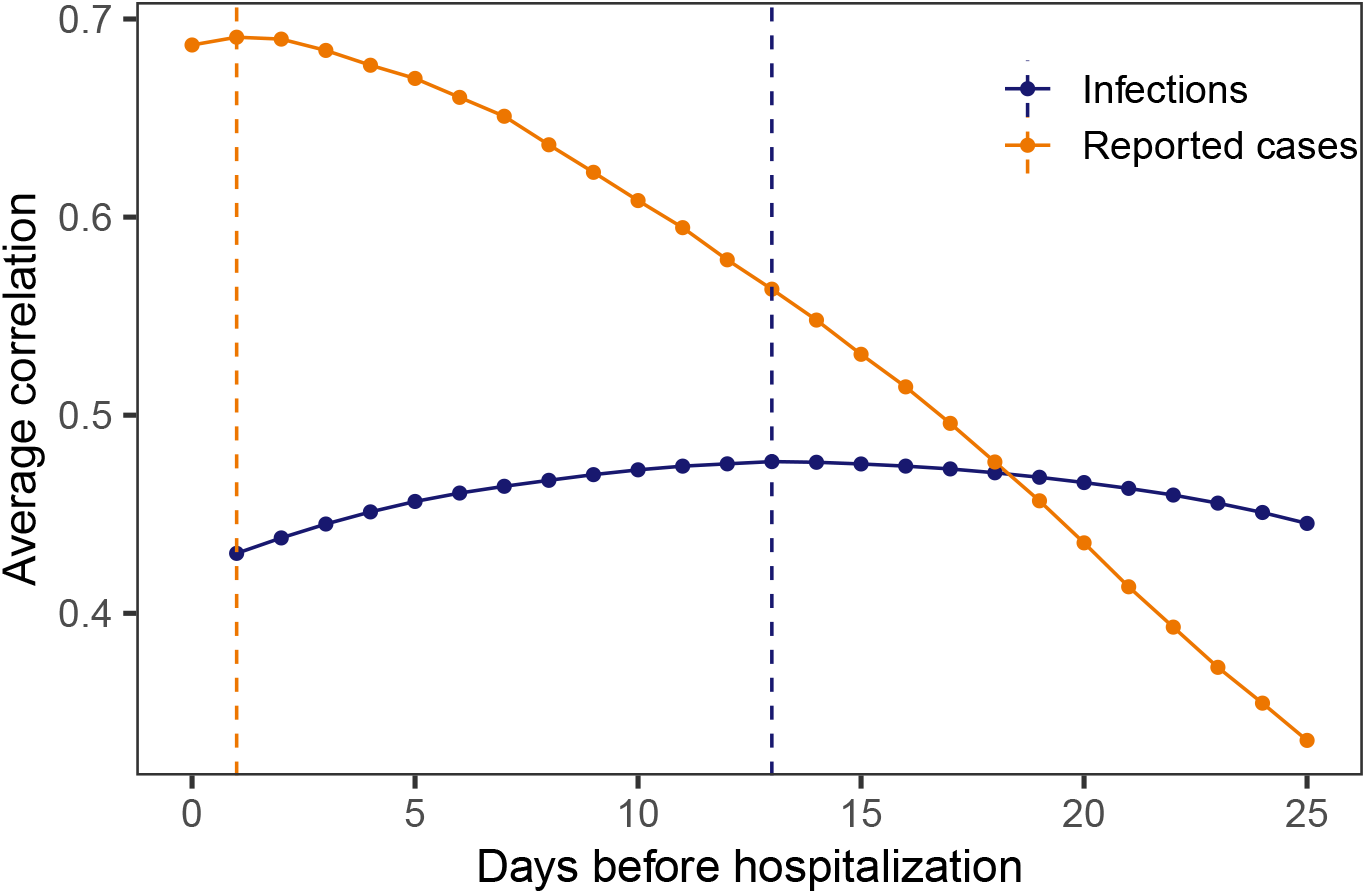
Spearman’s rank correlation between each of infections and cases with hospitalizations per 100,000. A rolling window of 61 days is applied before averaging across all states and times for each lag. The vertical dashed lines indicate the lags for which the highest average correlation is attained.

We compute the time-varying infection-hospitalization ratios (IHRs) for each state using a 13-day lag and case-hospitalization ratios (CHRs) with a 1-day lag for comparison Figure 7). Overall, the relationship between infections and hospitalizations is complex. It is characterized by intermittent spikes that punctuate longer periods where the IHRs are relatively stable, remaining below 0.1 hospitalizations per infection.

**Figure 7:**
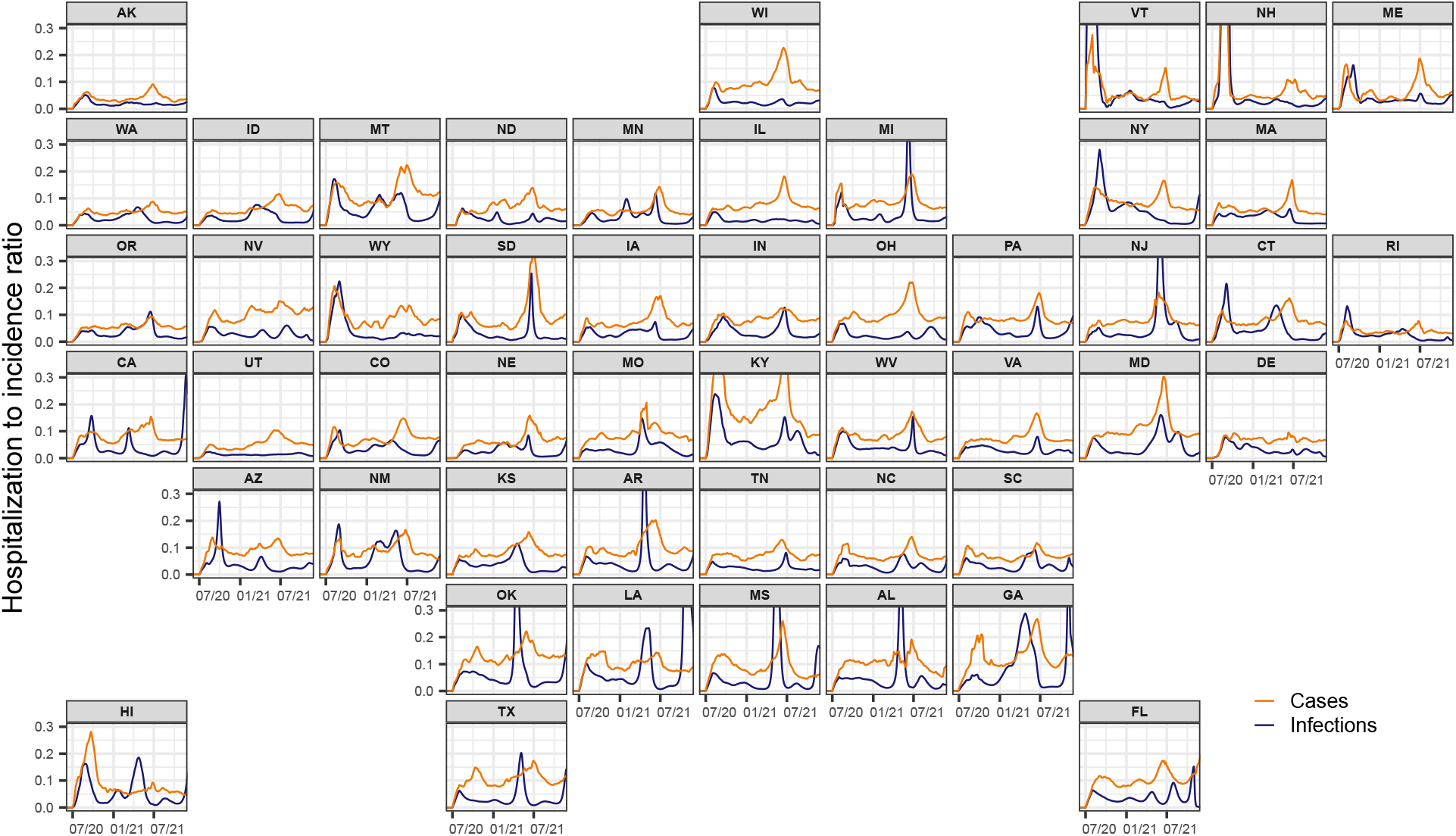
Time-varying IHR and CHR estimates for each state from June 1, 2020 to November 29, 2021, obtained using the respective correlation maximizing lag from Section 3.2. Note that the infection, case, and hospitalization counts are subject to a center-aligned 7-day average to remove spurious day of the week effects. Also note that the different starting points across states are due to the availability of the hospitalization data.

Both IHRs and CHRs exhibit similar spatiotemporal trends as those noted for infections. Namely, states that are proximate (for example, North and South Carolina) show similar temporal patterns in IHRs and CHRs. In addition, similar spikes are evident across many states during waves of infections that are driven by variants of concern. For example, many states exhibit a striking increase in hospitalizations in mid-2021, which coincides with the rapid takeover of the Delta variant (Hodcroft, 2021).

## 4 Discussion

We retrospectively estimated daily incident infections for each U.S. state over the period June 1, 2020 to November 29, 2021. Our estimates support the intuition that the pandemic impacted states earlier and at a larger scale than is indicated by reported cases. They also emphasize that using cases as a proxy for infections can lead to erroneous conclusions about trends in infections. More importantly, we observe outbreaks in infections that are missed from inspecting cases alone such as the Delta wave in New Jersey, Connecticut, and Maryland. These sorts of omissions serve to emphasize that cases paint an incomplete picture of the pandemic, especially when outbreaks are largely driven by unreported infections. Furthermore, since case reports generally follow symptom and infection onsets, cases have a built-in temporal bias. This is in addition to other biases from differences in reporting across states such as temporary bottlenecks due influxes of data or more persistent processing issues that increase the average time from case detection to report (Dunkel, 2020; Washington State Department of Health, 2020). Thus, while reported cases provide an indication of the trajectory of the pandemic, it is delayed and incomplete.

Our approach offers a number of advantages. By incorporating state-level case, line list, and variant circulation data, we are able to construct incubation and delay distributions that are spatiotemporally specific. Time-varying and state-specific seroprevalence data allows the reporting ratio estimates to similarly vary over space and time, a departure from existing work (Center for the Ecology of Infection Diseases, 2020; Unwin et al., 2020). Unlike previous approaches that use a single delay distribution to generate estimates for all states (Chitwood et al., 2022; Jahja et al., 2022; Miller et al., 2022), our work avoids this assumption of geographic invariance, an assumption that is far from realistic due to differences in the reporting pipelines, pandemic response, and variants in circulation, among other things. Similarly, prior methodology relies on only one incubation period distribution (Miller et al., 2022), whereas our method incorporates variant-specific incubation periods. This enhances our infection onset estimation by accounting for the differences across variants–specifically, that newer variants tend to have shorter incubation periods (Ogata et al., 2022; Tanaka et al., 2022; Wu et al., 2022).

Another limitation of previous approaches to estimate infections is that they often fail to account for reinfections. While reinfections constitute a small portion of the total infections until the arrival of high immune-escape variants (Omicron BA.1), disregarding them means that the infection-reporting ratio will tend to be underestimated with seroprevalence data alone. By accounting for reinfections as well as the waning of seropositivity, we more accurately estimate this ratio. However, future work could refine this analysis. Because the waning of immunity is likely to be variant-dependent (Pooley et al., 2023), our model’s single waning parameter would be more accurately estimated as a mixture of variant-specific parameters with weights determined by the proportion of the variants circulating.

We chose to end our analysis on November 29, 2021, for two main reasons. The first is that Omicron and subsequent variants come with substantial increases in the risk of reinfection in comparison to previous variants, likely due to increased immune escape (Eythorsson et al., 2022; Pulliam et al., 2022; Wei et al., 2024). Access to reinfection data that is representative of each location under study is paramount for extending the analysis. While it would be ideal to use the reinfection rates over time for each U.S. state, many states do not publicly report reinfection data over the entire time period under examination, if at all. The second reason is that the case-ascertainment ratio after December 2021 can no longer be estimated with seroprevalence data alone. Specifically, while most state-level data suggests that reinfections still account for less than 20% of reported cases during Omicron (Hawaii Department of Health, 2022; New York State Department of Health, 2023; Ruff et al., 2022; Washington State Department of Health, 2022), seropositivity rapidly reaches nearly 100% of the population. Therefore, alternative data sources for estimating the case-ascertainment ratio must be considered. For example, wastewater surveillance data may be complementary to seroprevalence data, especially when testing is low, or serve as a substitute when it is unavailable (McManus et al., 2023). An alternative approach could integrate surveillance streams from surveys, helplines, or medical records if they offer a sufficiently strong signal of the disease intensity over time (European Centre for Disease Prevention and Control, 2020; Reinhart et al., 2021).

Our work develops a deconvolution-based approach to inferring infection onset, combining available line list data with variant circulation estimates and literature derived incubation periods. This approach is complemented with the development of a model that incorporates waning detectable antibody levels and major seroprevalence surveys. The resulting infection estimates as well as their geospatial and temporal trends are strongly grounded in both data and statistical models.

These well-informed, localized estimates of COVID-19 infections provide a clear and comprehensive understanding of the pandemic’s progression over time. They contribute important information on the timing and magnitude of the disease burden for each location, and highlight trends that may not be visible from reported case data alone. Therefore, these infection estimates provide key information for the ongoing investigation on the true size and impact of the pandemic.

## Data availability

The required materials and code for reproducing all figures and the numerical results are available at https://github.com/cmu-delphi/latent-infections/.

## Acknowledgements

We would like to thank members of the Delphi research group for valuable feedback, and Change Healthcare and Optum/United Health Group for their invaluable data partnership and collaboration.

We gratefully acknowledge all data contributors, i.e., the Authors and their Originating laboratories responsible for obtaining the specimens, and their Submitting laboratories for generating the genetic sequence and metadata and sharing via the GISAID Initiative (Elbe and Buckland-Merrett, 2017), on which this research is based.

Any opinions, findings, and conclusions or recommendations expressed in this material are those of the authors and do not necessarily reflect the views of the National Science Foundation and the Centers for Disease Control and Prevention.

DJM and RJT were supported by Centers for Disease Control and Prevention (CDC) Grant No. 75D30123C15907. DJM and RL received support from the National Sciences and Engineering Research Council of Canada and the University of British Columbia. AS was supported by the Centers for Disease Control and Prevention and the National Science Foundation under Award No. 2223933 and 2333494.

