## Supplement for "Incident COVID-19 infections before Omicron in the U.S"

**Supplementary References** **15**

8 Lagged Spearman's correlation between the infection and hospitalization rates per 100,000 averaged for each lag across US states and days over June 1, 2020 to November 29, 2021, and taken over a rolling window of 61 days. The infection rates are based on the counts for the deconvolved case and infection estimates as well as the reported infections by symptom onset and when the report is symptom onset. Note that each such set of infection counts is subject to a center-aligned 7-day averaging to remove spurious day of the week effects. The dashed lines indicate the lags for which the highest average correlation is attained. . . . . . . . . . . . . . . . . . . . . . . . . . . . . . . . . . . 14

### List of Tables

1 Percent pairwise occurrence for the different permutations of events considered in the restricted CDC line list. The abbreviation IO stands for infection onset, SO is symptom onset, PS is positive specimen, and RE is report date. We consider a restricted set of permutations because we assume that IO must come first and that PS must precede report date for a case to be legitimate. Finally, the underlying assumption for the percent pairwise occurrence calculations is that the cases must have both elements present (not missing). . . . . . . . . 4

### 1 Supplementary Methods

#### A A general description and depiction of convolution

In general, the goal of convolution is to propagate the input signal forward in time using a probability distribution. In the 1D and discrete context, it is simply a rolling, weighted average of the past. So for an input sequence $\{x_t\}_{t=1}^n$ and time-constant weights $\{z(k)\}_{k=-\infty}^0$, the output sequence $\{y_t\}_{t=1}^n$ is given by

$$y_t = \sum_{s=0}^{t} z(s) x_{t-s}. \quad (1)$$

Figure 1 presents a depiction of the convolution procedure for an example signal $x_t$ (smoothed cases, orange line). Essentially, to push the cases forward in time, we take the appropriately aligned (forward-in-time) delay distribution $z(s)$ (blue shaded region) and convolve it with the smoothed cases signal counts by it to get the convolved estimates (blue line). This process is repeated as we march forward in time, as shown through the stop-motion panels, such that it eventually covers the entire line of cases. An important takeaway from this is that convolution is not the same as a simple shift of the data. A shift is a special case: when $z(d) = 1$ and $z(s) = 0$, $s \neq d$, we shift $x$ forward by $d$. Rather, convolution generally weights the entire past by non-zero probabilities. Deconvolution proceeds in the same fashion, but in the opposite direction, going backward in time and undoing the effect of a convolution.

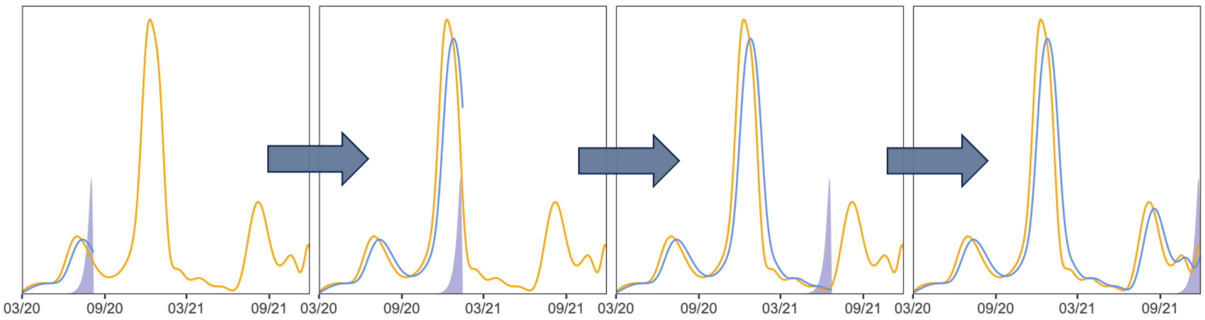

Figure 1: A general depiction of convolving smoothed cases (orange line) with the corresponding delay probabilities (shaded blue area) to get the convolved estimates (blue line) over four different times.

### B Additional details on the date fields in the CDC line list

Because the restricted CDC line list is updated monthly and cases may undergo revision, we use a single version of it that was released on June 6, 2022. We consider this version to be finalized in that it is well-beyond our study end date such that the dataset is unlikely to be subject to further significant revisions.

Table 1 presents the percent of pairwise occurrences for the different possible permutations of events in the line list. Essentially, most cases follow the idealized ordering shown by Figure 1 and so we adhere to this construction as much as possible. Unfortunately, the line list has significant missing data, notably with respect to our variables of interest. Approximately 62.3% of cases are missing the symptom onset date, 55.4% are missing positive specimen date, and 8.96% of cases are missing the report date. Furthermore, cases with missing report or positive specimen dates may be filled with their symptom onset date. Jahja et al. (2022) So it is possible that all three variables may have the same date for a case. However, we only actually deal with pairs of these events; we do not use all three at once in our construction of the delay distributions. Therefore, we restrict our investigation of coincident missingness to the possible pairs.

Due to the contamination in the zero delay cases (those whose symptom onset was used to fill missing positive specimen or report date, the true extent of which is unknown to us), we omit all cases where the positive specimen and report dates have zero delay from our analysis. We choose to allow for zero and negative delay for symptom onset to report because correspondence with the CDC confirms the possibility that a person could test positive before symptom onset and it is a reasonable ordering to expect if, for example, the individual is aware that they have been exposed to an infected individual.

| **Order of events** | **Percent pairwise occurrence** | **Handling** |
|---|---|---|
| IO $\rightarrow$ SO $\rightarrow$ PS $\rightarrow$ RE | PS $\geq$ SO: 97.1<br>PS = SO: 33.6<br>PS $>$RE: 1.74<br>PS = RE: 14.6 | This is the idealized order of events and so we built the current support sets for<br>SO $\rightarrow$ PS and PS $\rightarrow$ RE<br>delay distribution constructions around this such that IO comes first by construction,<br>SO typically precedes PS, but may be the same or come before, and RE comes after PS and SO |
| IO $\rightarrow$ PS $\rightarrow$ SO $\rightarrow$ RE | PS $<$SO: 2.91<br>SO $\leq$ RE: 99.3<br>SO $<$RE: 86.1 | Allowed for negative delays up to the largest non-outlier value for the 0.05 quantile of delay from PS to SO by state |
| IO $\rightarrow$ PS $\rightarrow$ RE $\rightarrow$ SO | RE $<$SO: 0.7<br>RE $<$PS: 1.7 | Nothing because current handling of the CDC of the line list ensures that the most concerning cases are handled where<br>SO = PO = RE, SO = RE and PO = RE |

Table 1: Percent pairwise occurrence for the different permutations of events considered in the restricted CDC line list. The abbreviation IO stands for infection onset, SO is symptom onset, PS is positive specimen, and RE is report date. We consider a restricted set of permutations because we assume that IO must come first and that PS must precede report date for a case to be legitimate. Finally, the underlying assumption for the percent pairwise occurrence calculations is that the cases must have both elements present (not missing).

The restricted CDC line list contains 74,849,225 cases (rows) in total compared to 84,714,805 cases reported by the JHU CSSE; that is, the line list is missing about 10 million cases. The extent that this issue impacts each state is shown in Figure 3, from which it is clear the fraction of missing cases is substantial for many states, often surpassing 50%. Jahja et al. (2022) In addition, the probability of being missing does not appear to be the same for states, so there is likely bias introduced from using the complete case line list data. We consider such bias to be unavoidable in our analysis due to a lack of alternative line list sources.

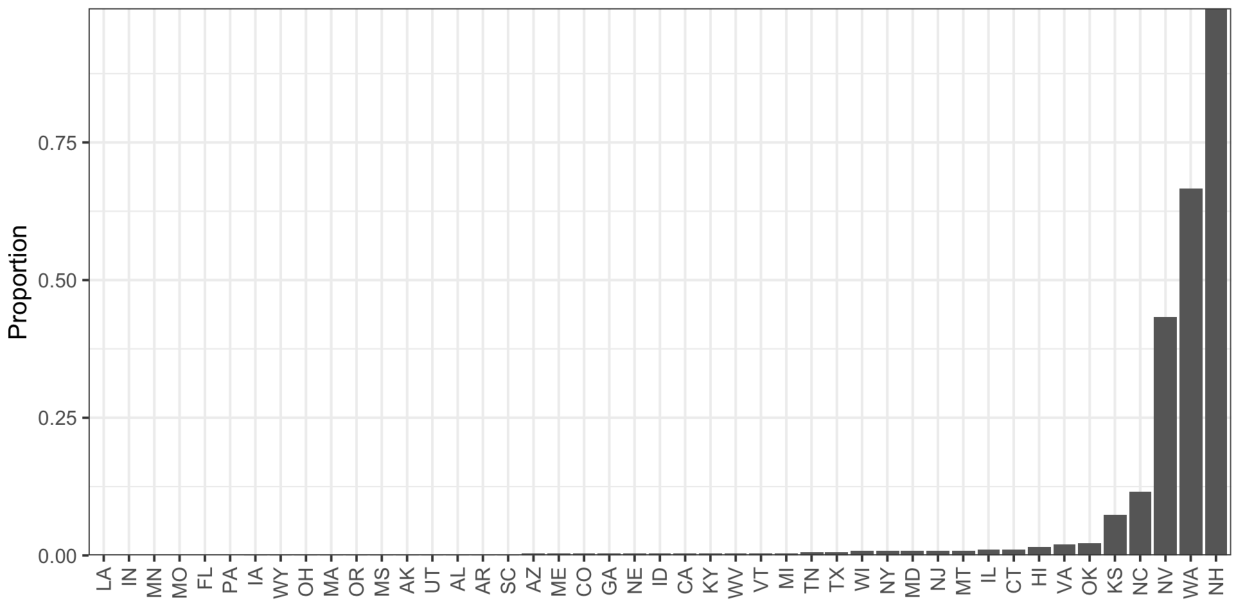

Figure 2: Proportion of complete cases with zero delay between positive specimen and report date in the restricted CDC line list dataset.

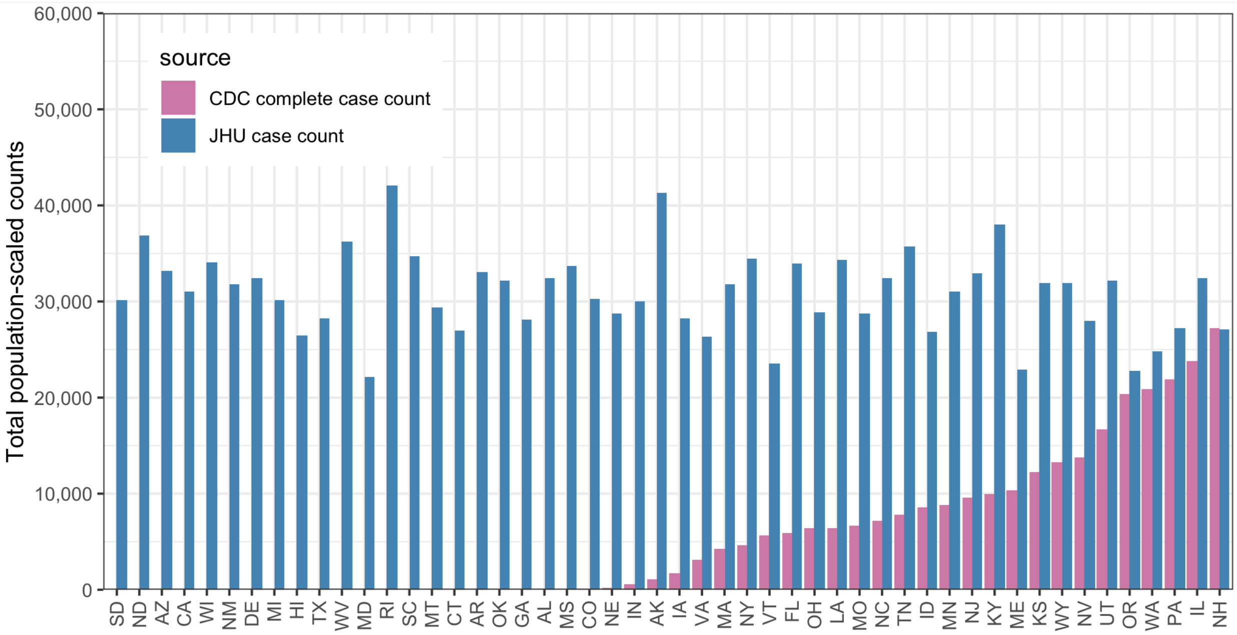

Figure 3: Complete case counts by state in the CDC line list versus the cumulative complete case counts from JHU CSSE as of June 6, 2022. All counts have been scaled by the 2022 state populations as of July 1, 2022 from (U.S. Census Bureau, Population Division, 2022).

#### C Additional details on delay distribution calculations

In the line list, we observe unusual spikes in reporting in 2020 in comparison to majority of 2021. When stratified by report date, a few states contribute unusually large case counts on isolated dates very late in the reporting process (often more than 100 days following specimen collection). These large accumulations of cases over time are likely due breakdowns of the reporting pipeline. Such anomalies are not likely to be reliable indicators of the delay from positive specimen to case report. Therefore, we prune these reporting backlogs systematically. The heuristic is to find large batches of cases that were all simultaneously reported on the same date with a lengthy delay (as would happen if a state "found" a tranche of previously unreported positive tests).

For each of three time intervals (delimited by July 16, 2020, October 16, 2020, and January 15, 2021), we apply the following pruning procedure:

Let $x_{\ell t}(d)$ be the number of cases in state $\ell$ with positive specimen collection date $t$ and delay $d$. For each $j = 1, 2, \ldots$, define

$$z_{\ell bj} = \sum_{d=50j}^{50(j+1)} \sum_t x_{\ell t}(d).$$

Then, locate the collection of potentially problematic combinations

$$\mathcal{H} = \{(\ell, b, j) : \log(z_{\ell bj}) > \text{median}_\ell(\log(z_{\ell bj})) + 1.5 \times \text{IQR}_\ell(\log(z_{\ell bj}))\}.$$

Remove any case reports from the line list in $\mathcal{H}$ where the total number of cases with the same report date exceeds 2000 if $j = 1$ or 500 otherwise.

Finally, in New Hampshire for a small handful of report dates, all cases reported by JHU appear in the CDC line list and all are recorded as having positive specimen collection date equal to the report date. The resulting estimate of the delay distribution (see Section 4.1) $\widetilde{p}_{\text{NH},t}(k)$ would be a point mass at $k = 0$ and the weight $\alpha_{\text{NH},t} = 1$ resulting $\widehat{\pi}_{\text{NH},t}(k)$ also being a point mass at $k = 0$. In this specific case, we force $\alpha_{\text{NH},t} = \min_{\ell \neq \text{NH}} \alpha_{\ell,t}$.

#### D Variant circulation proportions

To estimate the daily proportions of the variants circulating in each state, we obtain the GISAID genomic sequencing data from CoVariants.org. (Elbe and Buckland-Merrett, 2017; Hodcroft, 2021) These counts represent the total number of cases belonging to a particular variant using a sample of positive tests over a biweekly period. To estimate the population proportion of each variant, we apply multinomial logistic regression for the eight variant categories separately for each state. Multinomial logistic regression is a standard technique to model the frequency of SARS-CoV-2 variants. (Annavajhala et al., 2021; Figgins and Bedford, 2021; Obermeyer et al., 2022)

We let $V_{j\ell,t}$ to be the probability of a new cases at time $t$ in location $\ell$ corresponding to variant $j$. Let $v_{j\ell,t}$ be the analogous observed proportion. Then nonparametric multinomial logistic regression models the log odds as the system

$$\log\left(\frac{V_{j\ell,t}}{1-V_{j\ell,t}}\right) = \beta_{j\ell,0} + \beta_{j\ell,1}t + \beta_{j\ell,2}t^2 + \beta_{j\ell,3}t^3, \quad j = 1, \ldots J. \tag{2}$$

This is estimated along with a constraint to ensure that the estimated proportions will sum to 1 across all $J$ variants. The specification of the log odds as a third-order polynomial in time produces smoothness of the estimated proportions. Figure 4 shows the proportions by variant for California before (left) and after (right) the smoothing procedure.

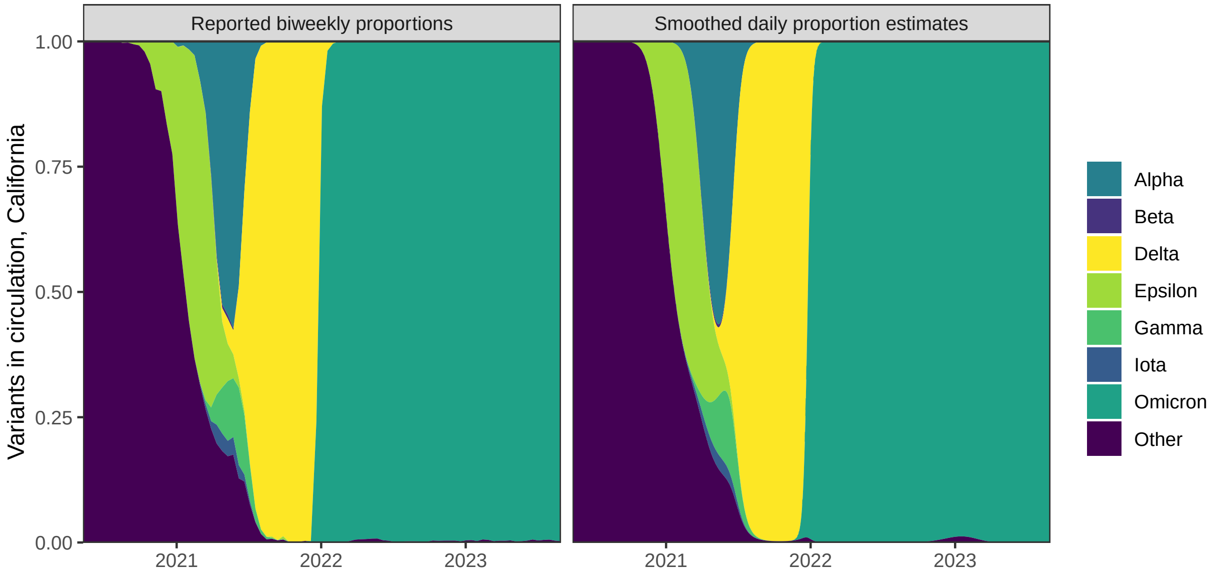

Figure 4: Left: Original biweekly proportions of the variants in circulation for California. Right: Daily proportions of the variants in circulation for California.

### E Constructing the delay from infection to test

The result of Step 1 (Section 4.1) is $\widehat{x}_{\ell,t}$, case estimates by positive specimen date for each state. To continue, pushing this back to infection estimates, we need the variant-specific delays from infection to positive specimen collection. As shown in Figure 1, this delay can be broken into two separate pieces: (1) the delay from infection to symptom onset, and (2) the delay from symptom onset to positive specimen collection. The first requires different methods and is specific to the variant causing the infection, while the second is estimable from the CDC line list.

#### E.1 Estimating the incubation period distributions

To account for the incubation period, the time between infection and symptom onset, we use estimates from the existing literature, modified slightly for coherence with each other: we model each incubation as a gamma distribution with different parameters. We focus on the following eight variants (shown in Figure 4, which saw significant circulation in one of the US states during our study period: Ancestral/Other, Alpha, Beta, Epsilon, Iota, Gamma, Delta, and Omicron. Alpha, Beta, Delta, Gamma, and Omicron are all variants of concern. World Health Organization (2021) while we include the Epsilon (California) and Iota (New York) variants because of large impact on those and neighbouring states. (Duerr et al., 2021; Yang et al., 2022)

The incubation period of the Ancestral variant has been modelled as a gamma distribution, Tindale et al. (2020) so we simply use the reported shape and scale parameters. For the Alpha, Beta, Gamma, Delta and Omicron variants, the mean and standard deviation are reported. (Grant et al., 2022; Ogata et al., 2022; Tanaka et al., 2022) Therefore, we use method of moments to match the mean and variance to estimate the gamma parameters, using the moment equations given in Section 4.1. Then, we discretize each resulting density shown in Figure 5 to the support set, which is taken to be from 1 and 21 days. This range assumes that symptoms require at least 1 day to develop Public Health Agency of Canada (2021) and that an asymptomatic infection will resolve within 21 days. (Cortes Martinez et al., 2022; Zaki and Mohamed, 2021)

We were unable to locate incubation period estimates for the geo-specific Epsilon and Iota variants, so we use the incubation period for Beta because Epsilon, Iota, and Beta are all children from the same parent in the phylogenetic tree of the Nextstrain Clades. Hodcroft (2021) All other circulating variants are grouped together with the Ancestral variant. There was little available sequencing data prior to Alpha-emergence, but unfortunately, later in the pandemic, it is impossible to separate Ancestral from other rare variants, though these also saw minimal circulation after the middle of 2021.

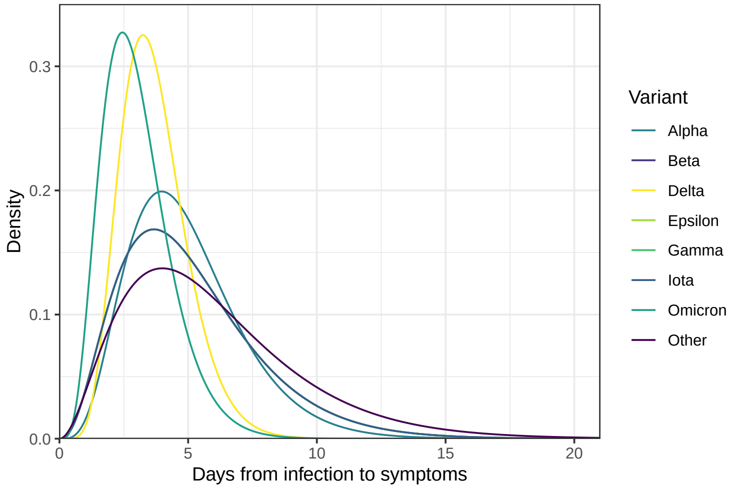

Figure 5: Gamma density for the incubation period of each of the eight variant categories. Note that the Ancestral variant uses reported shape and scale parameters. Tindale et al. (2020) while the remaining variants convert reported estimates for the mean and variance using the method of moments to produce the gamma parameters. (Grant et al., 2022; Ogata et al., 2022; Tanaka et al., 2022)

##### E.2 Estimating the delay distributions for symptom onset to positive specimen

Estimating the delay from symptom onset to positive specimen date follows a similar procedure as described in Section 4.1 with a minor adjustment. Here, we allow $k$ to range from $-3$ to 21 (rather than 1 to 60). These upper and lower bounds are based on the largest delay values for the state-wide 0.05 and 0.95 quantiles. The median delay is very short at approximately 2 days, and an asymptomatic individual may test positive following a known exposure, before the onset of symptoms. We show both types of delays for a sample of states over several dates in Figure 6. Unlike the delay from positive specimen collection to report, the delay from symptoms to positive specimen can conceivably be negative. The most obvious reason for this would be if a person knew they had been exposed to an infectious individual and so got tested prior to the development of symptoms. Required regular testing for jobs in health care settings, construction, or the film industry could also produce negative delays.

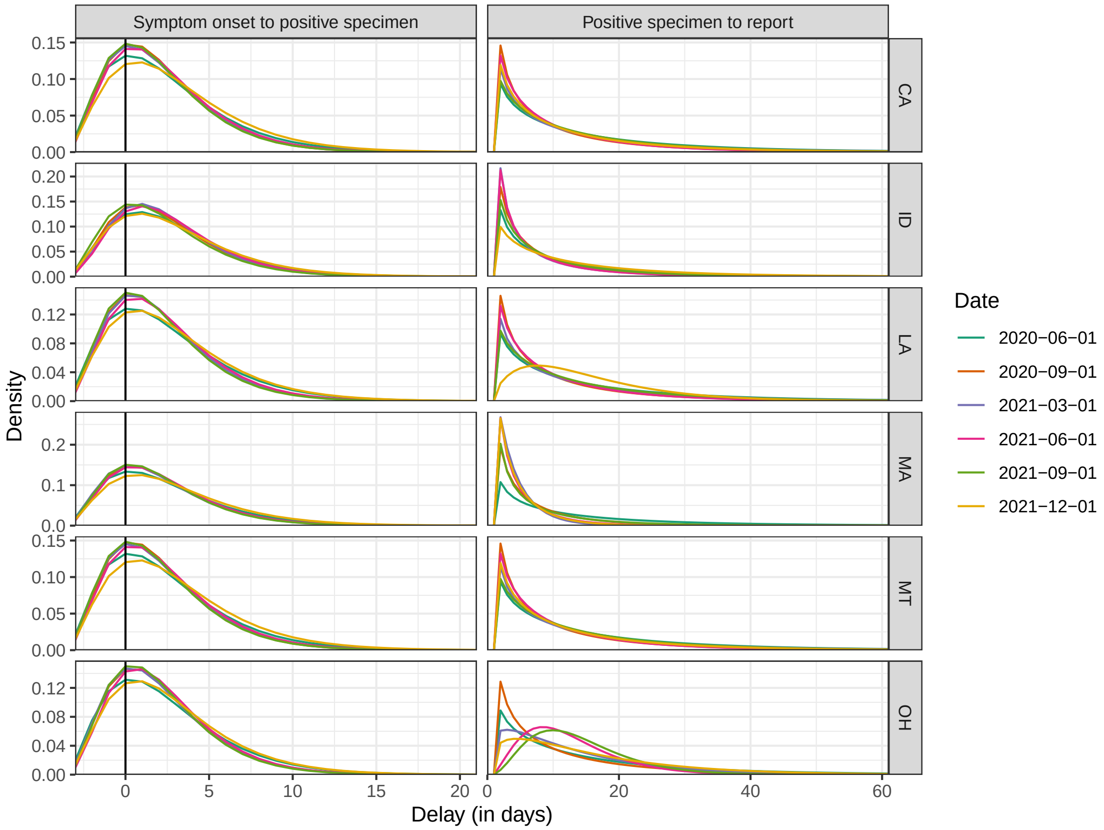

Figure 6: Depictions of the estimated delay from symptom onset to positive specimen date (left) and from positive specimen date to report date (right) for a sample of six states over several dates.

##### E.3 Details on constructing the infection-to-test distributions

Finally, to produce the delay from infection to positive specimen collection we convolve the variant-specific incubation periods from Section E.1, denoted by $\{i_j(k) : 0 < k \leq 21\}$ with the location-time-specific symptom-to-positive-test distributions from Section E.2, denoted by $\{q_{\ell,t}(k) : -3 \leq k \leq 21\}$. The convolution of these yields a distribution $\mathbf{q}_{\ell,t} * \mathbf{i}_j = \{\tau_{j\ell,t}(k) : -3 \leq k \leq 42\}$. Figure 6 shows the delays used for a sample of 6 states: symptom to positive specimen (left column) and positive specimen to report (right column). The convolution distribution $\tau_{j\ell,t}$ requires convolving the distribution in the left column with the variant-specific incubation periods shown in Figure 5.

#### F Details about seroprevalence data

We use two major contemporaneous surveys to estimate the proportion of the population with evidence of previous infection in each state over time: the 2020–2021 Blood Donor Seroprevalence Survey and the Nationwide Commercial Lab Seroprevalence Survey. (Centers for Disease Control and Prevention, 2021a, 2021b) In the former, the CDC collaborated with 17 blood collection organizations in the largest nationwide COVID-19 seroprevalence survey to date. Centers for Disease Control and Prevention (2021a) The blood donation samples were used to construct monthly seroprevalence estimates for nearly all states from July 2020 to December 2021. Jones et al. (2021) In the latter survey, the CDC collaborated with two private commercial laboratories to test blood samples from people that were in for routine or clinical management (presumably unrelated to COVID-19 Bajema et al. (2021)) for the antibodies to the virus. The resulting dataset contains seroprevalence estimates for a number of multi-week collection periods starting in July 2020 to February 2022.

Both datasets are based on repeated, cross-sectional studies that estimate the percentage of people who were previously infected with COVID-19 using the percentage of people from a convenience sample who had antibodies against the virus. (Bajema et al., 2021; Centers for Disease Control and Prevention, 2020; Jones et al., 2021) Adjustments were made in both for age and sex to account for the demographic differences between the sampled and the target populations. However, both datasets are incomplete and they differ in the number and the timing of the data points for each state (Figure 7). For example, in the commercial dataset, the last estimate for North Dakota is in September 2020. In the blood donor dataset, Arkansas does not have estimates available until October 2020.

A major difference in the structure of the two datasets is that the commercial dataset always has the seroprevalence estimates at the level of the state, while the blood donor dataset can either have estimates for the state or for multiple separate regions within the state. For the commercial dataset, we use the midpoint of the provided specimen collection date variable. For the blood donor dataset, we use the median donation date if the seroprevalence estimates are designated to be for entire state. If they are instead for regions in the state, since there is reliably one measurement per region per month, we aggregate the measurements into one per month per state by using a weighted average (to account for the given sample sizes of the regions). The median of the median dates is taken to be the date for the weighted average.

If there are multiple measurements in a week from a seroprevalence source, then the average is used.

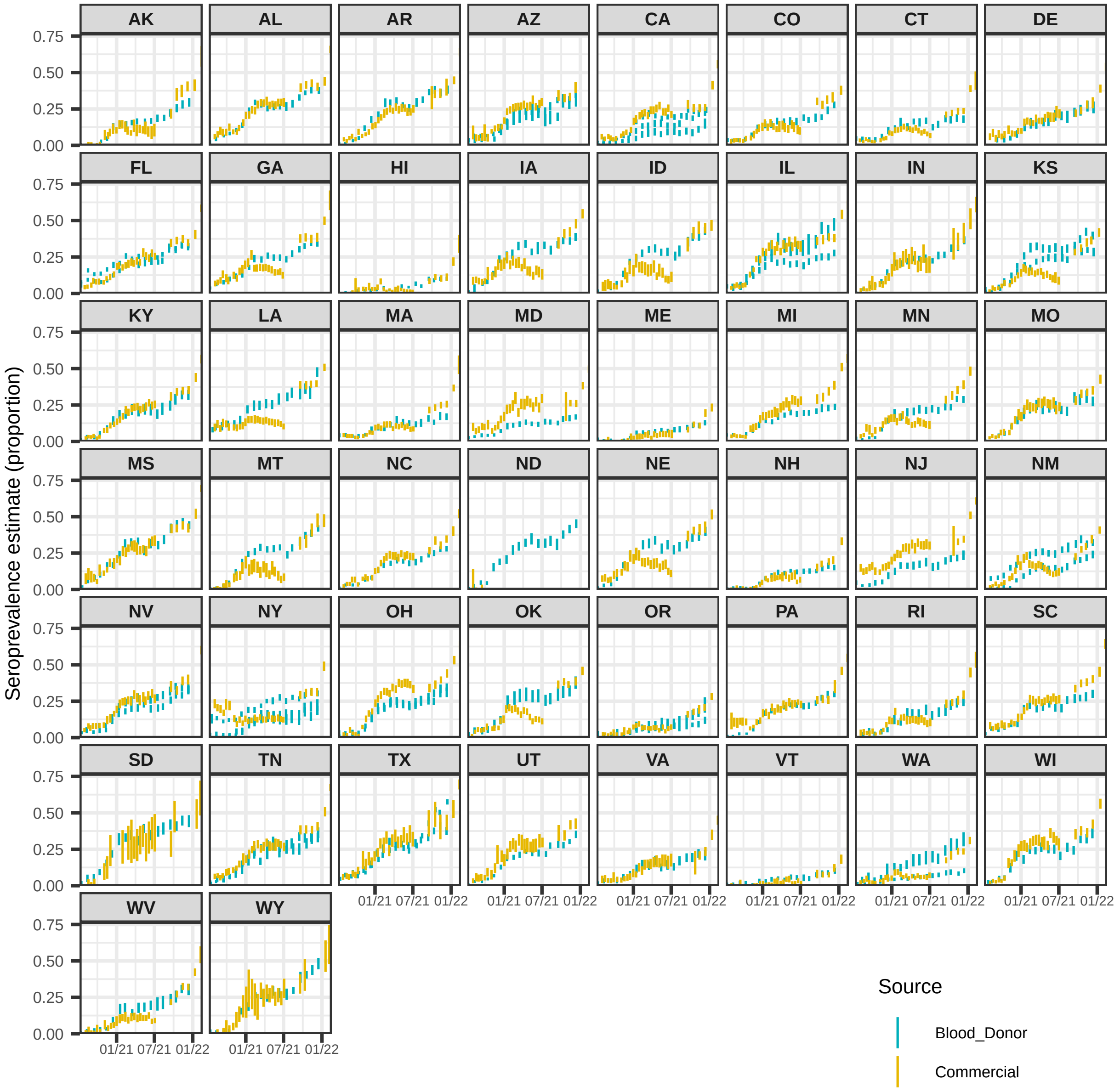

Figure 7: A comparison of the seroprevalence estimates from the Commercial Lab Seroprevalence Survey dataset (yellow) and the 2020–2021 Blood Donor Seroprevalence Survey dataset (blue). Note that the maximum and the minimum of the line ranges are the provided 95% confidence interval bounds to give a rough indication of uncertainty.

### G State space representation of the antibody prevalence model

The antibody prevalence model can be expressed as a linear Gaussian state space model. (Durbin and Koopman, 2012; Helske, 2017) For $m = 1, \ldots, M$, let $\alpha_m$ be a vector of latent state processes at time $m$ and $y_m$ be a vector of observations at time $m$. The form of the (general) linear Gaussian state space model is

$$y_m = Z\alpha_m + \sigma_r^2 \epsilon_m, \qquad \epsilon_m \sim N(0, H_m) \tag{3}$$

$$\alpha_{m+1} = T_m \alpha_m + R\eta_m, \quad \eta_m \sim N(0, Q) \tag{4}$$

where $\alpha_1 \sim N(a_1, P_1)$ and $\epsilon_m$ and $\eta_m$ are mutually and serially independent.

To express the antibody prevalence model in state space form, we define the components in Equations (3) and (4) as follows (omitting the location subscript for simplicity):

$$R = \begin{bmatrix} 1 & 0 \\ 0 & 1 \\ 0 & 0 \\ 0 & 0 \end{bmatrix} \qquad Z = \begin{bmatrix} 1 & 0 & 0 & 0 \\ 0 & 1 & 0 & 0 \end{bmatrix} \qquad H_m = \begin{bmatrix} w_m^1 \sigma_r^2 & 0 \\ 0 & w_m^2 \sigma_r^2 \end{bmatrix}$$

$$\alpha_m = \begin{bmatrix} s_m \\ a_m \\ a_{m-1} \\ a_{m-2} \end{bmatrix} \qquad T_m = \begin{bmatrix} (1-\gamma) & \widehat{u}_m^{\Sigma}(1-z_m) & 0 & 0 \\ 0 & 3 & -3 & 1 \\ 0 & 1 & 0 & 0 \\ 0 & 0 & 1 & 0 \end{bmatrix} \qquad Q = \begin{bmatrix} \sigma_\epsilon^2 & 0 \\ 0 & \sigma_\eta^2 \end{bmatrix}$$

$$a_1 = \begin{bmatrix} \tilde{s}_1 \\ \tilde{a}_1 \\ \tilde{a}_1 \\ \tilde{a}_1 \end{bmatrix} \qquad P_1 = \begin{bmatrix} \sigma_{\tilde{s}_1}^2 & 0 & 0 & 0 \\ 0 & \sigma_{\tilde{a}_1}^2 & 0 & 0 \\ 0 & 0 & \sigma_{\tilde{a}_1}^2 & 0 \\ 0 & 0 & 0 & \sigma_{\tilde{a}_1}^2 \end{bmatrix}$$

where $\sigma_r^2$ is the variance of observations, $\sigma_\epsilon^2$ is the variance of the seroprevalence estimates, $\sigma_\eta^2$ is the trend variance, and $\widehat{u}_m^{\Sigma}$ denotes the new deconvolved cases between $m$ and $m+1$. Because the inverse reporting ratios should be more variable than the seroprevalence estimates, we enforce that the estimate of $\sigma_\eta^2$ is a multiple of $\sigma_\epsilon^2$.

Finally, $w_m^1$ and $w_m^2$ are the time-varying inverse variance weights computed from the commercial and blood donor datasets, respectively. For each source, we compute the weights for the observed seroprevalence estimates using the formula for the standard error of a proportion. These weights are then re-scaled to sum to the number of observed seroprevalence measurements for the source. Finally, the ratio of the average observed weights from the two sources is used to scale all weights relative the commercial source. This transformation is done purely for computational purposes to make estimation of the unknown parameters easier.

The prior distribution for $\alpha_1$ is estimated using both data-driven constraints and externally sourced information. The initial value of the seroprevalence component, $\tilde{s}_1$ is the average of the initial seroprevalence measurements from each source rounded down to two decimal places,

The corresponding initial variance estimate, $\sigma^2_{\tilde{s}_1}$, is taken to be the mean of the standard errors of the two seroprevalence estimates. For the initial values of the trend components, we use the inverse of the ascertainment ratio estimate for each as of June 1, 2020 from Table 1 in Unwin et al. (2020): the mean for $\tilde{a}_1$ and half the width of the confidence interval converted to the inverse for $\sigma^2_{\tilde{a}_1}$.

The initial $\sigma^2_r$ is taken to be the average of the estimated variances from the observed seroprevalence measurements regressed linearly on time. The initial value of the multiplier is set to be 100 for all states. The $\sigma^2_\epsilon$ and $\gamma$ values are estimated separately for each state, then fixed to their averages on the log-scale.

Following maximum likelihood estimation of remaining parameters we use the Kalman filter to obtain the smoothed point estimates and variances of the weekly inverse reporting ratios. We use forward and backward extrapolation to extend these estimates outside of the observed seroprevalence range Durbin and Koopman (2012) followed by linear interpolation to produce daily values. We then multiply these by the corresponding deconvolved case estimates before converting to per-capita values. Annual estimates of the state populations as of July 1 of 2020 are taken from the Dec. 2022 press release from the US Census Bureau. U.S. Census Bureau, Population Division (2022)

#### H Ablation analysis of infection-hospitalization correlations

In this ablation study, the lagged correlation is re-computed by using the following infection estimates: 1. Those from the deconvolution procedure under the assumption that the infection onset is the same as the positive specimen date (i.e., excluding the positive specimen to infection onset data and deconvolution); 2. Those from the deconvolution procedure under the assumption that the infection onset is the same as the symptom onset date (excluding the incubation period data); 3. Those from the deconvolution procedure when utilizing all incubation period and delay data (the deconvolved case estimates); 4. Those from applying the antibody prevalence model to produce estimates for both the reported and the unreported cases (the infection estimates).

The results of this study are shown in Figure 8. From this, we can see that the deconvolved case and infection estimates from the intermediate steps are all leading indicators of hospitalizations. However, the degree that each set of estimates leads hospitalizations depends on its location in the sequence of deconvolution steps and how close the estimates are to infection onset. For example, the deconvolved cases by positive specimen date tend to precede hospitalizations by about 11 days, while those for the subsequent step indicate that the deconvolved cases by symptom onset tend to precede hospitalizations by a longer time of 13 days. Finally, after adding the variant-specific incubation period data into the deconvolution and obtaining the deconvolved case estimates, we can observe that the reported infections precede hospitalizations by about 19 days.

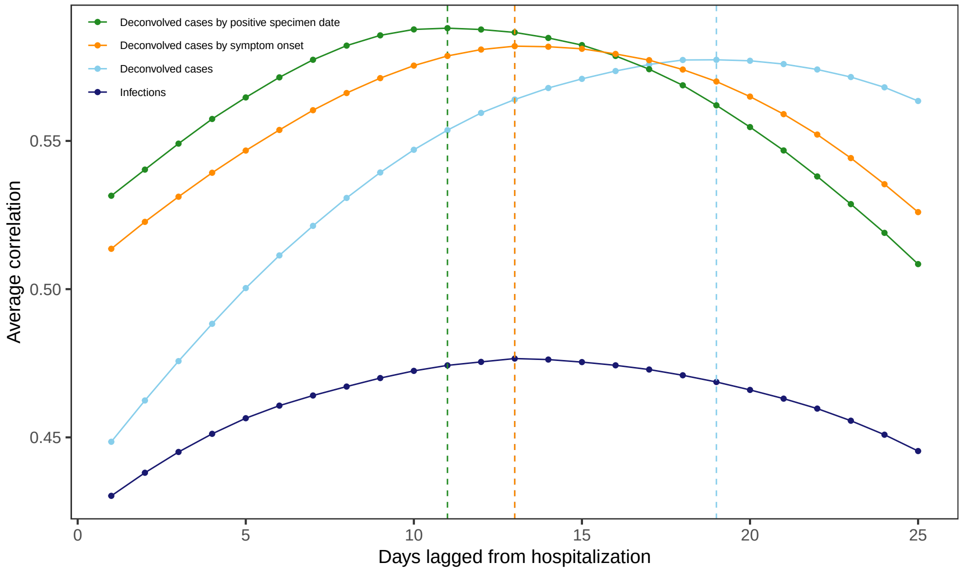

Figure 8: Lagged Spearman's correlation between the infection and hospitalization rates per 100,000 averaged for each lag across US states and days over June 1, 2020 to November 29, 2021, and taken over a rolling window of 61 days. The infection rates are based on the counts for the deconvolved case and infection estimates as well as the reported infections by symptom onset and when the report is symptom onset. Note that each such set of infection counts is subject to a center-aligned 7-day averaging to remove spurious day of the week effects. The dashed lines indicate the lags for which the highest average correlation is attained.

Centers for Disease Control and Prevention, 2021b. Nationwide commercial laboratory seroprevalence survey.

Centers for Disease Control and Prevention, 2020. COVID Data Tracker.

Cortes Martinez, J., Pak, D., Abelenda-Alonso, G., Langohr, K., Ning, J., Rombauts, A., Colom, M., Shen, Y., Gomez Melis, G., 2022. SARS-CoV-2 incubation period according to vaccination status during the fifth COVID-19 wave in a tertiary-care center in Spain: A cohort study. BMC Infectious Diseases 22, 1–7.

Duerr, R., Dimartino, D., Marier, C., Zappile, P., Wang, G., Lighter, J., Elbel, B., Troxel, A.B., Heguy, A., others, 2021. Dominance of Alpha and Iota variants in SARS-CoV-2 vaccine breakthrough infections in New York City. The Journal of Clinical Investigation 131, e152702.

Durbin, J., Koopman, S.J., 2012. Time series analysis by state space methods. OUP Oxford.

Elbe, S., Buckland-Merrett, G., 2017. Data, disease and diplomacy: GISAID's innovative contribution to global health. Global Challenges 1, 33–46.

Figgins, M.D., Bedford, T., 2021. SARS-CoV-2 variant dynamics across US states show consistent differences in effective reproduction numbers. MedRxiv 2021–12.

Grant, R., Charmet, T., Schaeffer, L., Galmiche, S., Madec, Y., Von Platen, C., Chény, O., Omar, F., David, C., Rogoff, A., others, 2022. Impact of SARS-CoV-2 Delta variant on incubation, transmission settings and vaccine effectiveness: Results from a nationwide case-control study in France. The Lancet Regional Health–Europe 13, 100278.

Helske, J., 2017. KFAS: Exponential family state space models in R. Journal of Statistical Software 78, 1–39.

Hodcroft, E., 2021. CoVariants: SARS-CoV-2 mutations and variants of interest.

Jahja, M., Chin, A., Tibshirani, R.J., 2022. Real-time estimation of COVID-19 infections: Deconvolution and sensor fusion. Statistical Science 37, 207–228.

Jones, J.M., Stone, M., Sulaeman, H., Fink, R.V., Dave, H., Levy, M.E., Di Germanio, C., Green, V., Notari, E., Saa, P., others, 2021. Estimated US infection-and vaccine-induced SARS-CoV-2 seroprevalence based on blood donations, July 2020-May 2021. JAMA 326, 1400–1409.

Obermeyer, F., Jankowiak, M., Barkas, N., Schaffner, S.F., Pyle, J.D., Yurkovetskiy, L., Bosso, M., Park, D.J., Babadi, M., MacInnis, B.L., others, 2022. Analysis of 6.4 million SARS-CoV-2 genomes identifies mutations associated with fitness. Science 376, 1327–1332.

Ogata, T., Tanaka, H., Irie, F., Hirayama, A., Takahashi, Y., 2022. Shorter incubation period among unvaccinated delta variant coronavirus disease 2019 patients in Japan. International Journal of Environmental Research and Public Health 19, 1127.

Public Health Agency of Canada, 2021. COVID-19 for health professionals: transmission.

Tanaka, H., Ogata, T., Shibata, T., Nagai, H., Takahashi, Y., Kinoshita, M., Matsubayashi, K., Hattori, S., Taniguchi, C., 2022. Shorter incubation period among COVID-19 cases with the BA. 1 Omicron variant. International Journal of Environmental Research and Public Health 19, 6330.

Tindale, L.C., Stockdale, J.E., Coombe, M., Garlock, E.S., Lau, W.Y.V., Saraswat, M., Zhang, L., Chen, D., Wallinga, J., Colijn, C., 2020. Evidence for transmission of COVID-19 prior to symptom onset. eLife 9, e57149.

Unwin, H.J.T., Mishra, S., Bradley, V.C., Gandy, A., Mellan, T.A., Coupland, H., Ish-Horowicz, J., Vollmer, M.A.C., Whittaker, C., Filippi, S.L., others, 2020. State-level tracking of COVID-19 in the United States. Nature Communications 11, 6189.

U.S. Census Bureau, Population Division, 2022. Annual estimates of the resident population for the United States, regions, states, District of Columbia, and Puerto Rico: April 1, 2020 to july 1, 2022.

World Health Organization, 2021. Tracking SARS-CoV-2 variants.

Yang, S., Hemarajata, P., Hilt, E.E., Price, T.K., Garner, O.B., Green, N.M., 2022. Investigation of SARS-CoV-2 Epsilon variant and hospitalization status by genomic surveillance in a single large health system during the 2020-2021 winter surge in Southern California. American Journal of Clinical Pathology 157, 649–652.

Zaki, N., Mohamed, E.A., 2021. The estimations of the COVID-19 incubation period: A scoping reviews of the literature. Journal of Infection and Public Health 14, 638–646.
